# DETERMINANTS OF PRETERM BIRTH IN PUBLIC HOSPITALS OF HORRO GUDURU WOLLEGA ZONE, WESTERN ETHIOPIA: UNMATCHED CASE CONTROL STUDY

**DOI:** 10.1101/2022.11.11.22282209

**Authors:** Warkisa Bayisa Duresa, Emiru Merdassa Atomssa, Bizuneh Wakuma, Worku Etafa Ebi

## Abstract

**Background:** Preterm birth is one of a public health issue worldwide. It is a single most important cause of death in the first month of life and second leading cause of death in children aged less than five years. The cause of preterm birth is multifactorial and not well identified. Therefore, the aim of this study was to identify determinants of preterm birth in public hospitals of Horro Guduru Wallaga zone, Western Ethiopia.

**Methods:** Hospital-based unmatched case-control study design was carried out in public hospitals of Horro Guduru Wallaga zone which involved 78 cases and 155 controls. Data were collected using pre-tested questionnaires from three hospitals. Data were entered in to EpiData and exported to SPSS version 25 for analysis. Frequency and proportion were computed to summarize the data. Ethical approval was made by Wollega University Research Ethical Review Committee before the study was started. Multivariable binary logistic regression analysis was used to determine the association of predictor and response variable at P < 0.05. Adjusted odds ratio with 95% CI was used to show the strength of association between predictors and outcome variables.

**Results:** Out of 233 mothers, 231(78 cases and 153 controls) were participated with response rate, 99.14%. The result of this study showed that not attending ANC (AOR= 4.61, 95%CI; 1.54, 13.79), history of abortion(AOR =3.88, 95% CI; 1.62, 9.30), Premature rupture of the membrane (AOR=3.91,95% CI;1.15,13.25), Sexually transmitted illness (AOR=3.51,95% CI; 1.26,9.76) and physical violence (AOR=2.78, 95%CI;1.19,6.52) had significant association with preterm birth.

**Conclusions:** The result of this study showed that not attending antenatal care, history of abortion, premature rupture of membrane, sexually transmitted illness and physical violence identified as potential risk factors of preterm birth. Therefore, it is important to encourage pregnant mothers to have optimum antenatal care follow-up. Healthcare providers should also focus on screening and counseling pregnant mother on obstetric complications and limiting physical violence.

## INTRODUCTION

The World Health Organization define preterm birth as any birth before 37 completed weeks of gestation since the first day of the woman’s last menstrual period. It is further divided into extremely preterm (<28 weeks), very preterm (28–<32weeks) and moderate or late preterm (32–<37weeks of gestation). Most preterm births happen spontaneously, but some are due to early induction of labor or caesarean birth, whether for medical or non-medical reasons(1).

Preterm birth is public health issue worldwide with the leading cause of death among neonates and the second leading cause of death among less than five children. Globally, almost 15 million babies are born too soon; for instance, more than one in ten newborns born too soon. Approximately, 90% of preterm birth is found in low and middle-income countries; whereas about 80% of preterm births occur in Sub-Sahara Africa and South Asia(2).

The burden of preterm birth is truly a global problem, not only confined to developing countries; for instance, United States and Brazil ranks among the top 10 countries. The magnitude of preterm birth were 10.2% in United states, 13.6% in India, 11.2% in Brazil, 19.1% in Bangladesh, 6.9% in China, 10.4% in Indonesia and 8.4% in Pakistan,16.6% in Tanzania,11.4% in Nigeria(3, 4) and 16.30% in Malawi(5), 20.20% in Kenya(6) and 12% in Ethiopia(7). Moreover, in Ethiopia the rate of preterm birth have significant regional disparities; that is 4.4% in Gondar(8), 35% in Dessie(9), 15.5% in Butajira(10),13.3% in Central zone Tigray(11), 16.9% in shire Tigray(12), 12.3% of Sumale(13) and 25.9% in Jimma(14).

Globally, more than 3.1million children death occur each year, and about 1.1million children mortality occur due to their prematurity(15). Thus, preterm birth is the leading cause of neonatal mortality, accounting 35% of neonatal deaths and the second most common cause of mortality next to pneumonia among children <5years, which is 18% of under five children death(2, 15). In developing countries, the risk of dying from preterm birth is 10times higher, and more than three fourth of perinatal mortality and more than half of long-term morbidity occurs due to their prematurity. Furthermore, prematurity has harmful consequences on individual, families and community (2, 16, 17).

Even though efforts have made on preterm prevention, diagnosis and intervention by Sustainable development goal which target “end all preventable neonatal and child death 2030" worldwide; the rate of neonatal and child death was still higher(18). Likewise, Ethiopia has been making a progress change, achieving most of national and global health indicators through Sustainable development goal, health sector transformation plan (HSTP) and Integrated Management of Newborn and Childhood Illnesses (IMNCI) standards to reduce the major cause of death in children less than five years including prematurity(19).

Moreover, currently, there is strong leadership, coordination, intensive health system investment and community involvement to promote maternal and child health. However, preterm birth is still one of the leading causes of childhood death that needs further strong study to identify potential determinants prematurity. In some parts of Ethiopia, studies show that preterm birth is the most critical and multifactorial with unknown specific risk factors. In addition, previous studies less emphasized on determinant factors, used secondary data and not addressed risk factors like partner violence, nutritional factor, alcohol and smoking. Moreover, there is very little information on determinants of preterm and no related study conducted in this area. The scarcity of available data on this topic in the study area has limited awareness on prevention and intervention strategies in line with Sustainable development goal. In this context, this study aimed to identify further determinants of preterm birth in public hospitals of HGW zone, Western Ethiopia.

## MATERIALS AND METHODS

Hospital-based unmatched case-control study was conducted public hospitals of Horro Guduru Wollega zone, western Ethiopia. These Hospitals: Shambu hospital, Guduru hospital and Abe dongoro hospital. The study was conducted from January 01, 2022 to April 30, 2022. The source population for this study was all mothers of newborn who gave birth in public hospitals of Horro Guduru zone and the study population was all mothers of newborn who gave birth in public hospitals of Horro Guduru zone during the study period, whereas, the study participants are all consecutively selected mother who gave birth in public hospitals of HGW zone, Western Ethiopia during the study period.

Inclusion criteria for Cases were all mothers of new preterm neonate (mothers who gave birth between 28 and 36 completed weeks of gestational) and controls were all selected mothers of new term neonate (mothers who gave birth at/after 37 weeks of gestational) in the public hospitals of HGW zone during the study period.

However, all mothers immediately referred out, unable to communicate, mothers who had stillbirth, neonates of maternal death, mothers who gave birth after 42 weeks of gestation, mothers didn’t know their last menstrual period, or had not taken early ultrasound assessment, and mother of newborn revisit the hospital during data collection period were excluded.

The sample size was calculated using Epi Info version 7 based on the assumptions 95% two-sided Confidence level, 80% power and Ratio of controls to cases was 2:1 and from previous information, proportion of maternal history of Anemia among control was 7.6% and odds ratio of 3.24. The calculated sample size was 233 (78 cases and 155 controls). Those women fulfilling inclusion criteria were recruited until the predetermined sample size was satisfied. For each case two consecutive controls were selected; while the selected mother did not fit the criteria, the next eligible mother was selected.

A structured questionnaire was developed to assess socio-demographic, lifestyle and behavioral factors, nutritional factors, medical and obstetric factors, fetal factors and Intimate partner violence among case and control mothers of preterm neonates gave birth in public hospitals. The questionnaire was first prepared in English version, and translated to ‘*Afaan Oromoo*’ version, which is the official working language of the region and then re-translated back to English, by language translators who were blind to the original questionnaire, to maintain consistency of the variables under question. The questionnaire was pre-tested on 10% of total sample size of study at Gedo Hospital and some adjustment was done to ensure the quality based on pretest result before actual data collection. Four trained midwives and two trained nurses recruited and collected the data, and three Master of public health experts supervised the data collection process. Two days training was provided to data collectors and supervisors on objectives, ethical issue, measurement scales, allowable criteria, how to complete the questionnaire and similar approach to the study subjects by experts on the area and principal investigator.

Data were collected through face to face interview. Collected data were checked daily by supervisor in each hospital for its completeness, clarity and consistency. Then, the collected data was reviewed and checked for completeness by supervisors and investigator. EpiData version 3.1 was used for data entry, and exported to SPSS version 25 for cleaning and further analyses.

Bivariable binary logistic regression analysis was used to select unadjusted effect of each independent variable on dependent variable, and variables with P-value <0.25 identified as a candidate variable for multivariable analysis.

The strength of association between dependent variable and independent variables was expressed using odds ratio (OR) through 95% confidence interval. Finally, multivariable binary logistic regression analysis was done to identify independent effect of each independent variable on preterm birth by accounting the effect of others. Multicollinearity assessment was conducted using the means of variance inflation factor as a post-estimation procedure following regression analysis. All covariates having value of Variance inflation factor (VIF) <10 were tolerable, while the presence of collinearity begin to be observed if VIF >10. The Hosmer-Lemshow goodness of fit test was carried out and P-value > 0.05 ensures the model adequately fitted the data and ROC curve was plotted for the fitted model and ROC with area under the curve of 0.70 and above were considered as acceptable discrimination. Results of Multivariable were interpreted with adjusted odds ratios (AOR) with 95% confidence intervals (95%CI) and variables with P-value less than 0.05 on multivariable binary logistic regressions were considered as statistically significant.

The study was approved by Wallaga University research ethical review committee (WURERC) on December 30, 2022 with Ref No.WU/RD/497/2022. Formal letters of co-operation were written to all selected hospitals and permission was secured at all levels. Informed consent was obtained from each study participant. The name or any identification of participant was not considered and privacy of each respondent was maintained throughout the research. The respondents have told that their participation was voluntary and had the right to withdraw from the study, even refuse at any time. The filled questionnaires were kept under secured custody of the corresponding author and entered into password secured computer. The purpose and process of the study was explained to all participants and concerned bodies.

## RESULTS

Out of 233 eligible mothers of newborn observation, 231 mothers (78 cases and 153 controls) were participated in the study with a response rate of 99.14%.

### Socio-demographic characteristics of study participants

Out of 231 mothers participated in the study; nearly three forth of mothers were in 25-34 age groups. The mean age of mothers among cases were 30.14 years (SD ±4.29), whereas the mean age of mothers in control was 30.02 years (SD± 4.35). About 43(55.1%) of cases were protestant Christianity follower, whereas 95(62.1%) of mothers in control group. About 55(70.5%) of mothers among case lives in rural setting, whereas 84(54.9%) among control group lives in the rural area. Among cases, about 71(91%) of mothers and 145(94.8%) of control were married, whereas 41(52.6%) of cases and 53(34.6%) of controls were unable to read and write. About nine (11.5%) of cases and 22(14.4%) of mothers among controls had greater than six members in family size (**Table 1**).

**Table 1:** Sociodemographic characteristics of mothers who gave birth in public hospitals of HGW zone, 2022 (n=231; case: 78 and control: 153)

| Socio demographic characteristics |  | Case, n (%) | Control, n (%) | Total, n (%) |
| --- | --- | --- | --- | --- |
| Age | 15-24 | 5(6.4) | 9(5.9) | 14(6.1) |
|  | 25-34 | 58(74.4) | 115(75.2) | 173(74.9) |
|  | 35-49 | 15(19.2) | 29(19.0) | 44(19.0) |
| Residence | Urban | 23 (29.5) | 69 (45.1) | 92 (39.8) |
|  | Rural | 55(70.5) | 84(54.9) | 139(60.2) |
| Marital Status | Married | 71(91.0) | 145(94.8) | 216(93.5) |
|  | Others* | 7(9.0) | 8(5.2) | 15(6.5) |
| Education level | No formal Education | 57(73.1) | 95(62.1) | 152( 65.8) |
|  | Formal Education | 21(26.7) | 58(37.9) | 79(34.2) |
| Religion | Degree and Above | 1(1.3) | 16(10.5) | 17(7.4) |
|  | Protestant | 43(55.1) | 95(62.1) | 138(59.7) |
|  | Orthodox | 23(29.5) | 41(26.8) | 64(27.7) |
|  | Muslim | 10(12.8) | 14(9.2) | 24(10.4) |
|  | Waaqeffataa | 2(2.6) | 3(2.0) | 5(2.2) |
| Occupational status | House wife | 46(59.0) | 79(51.6) | 125(54.1) |
|  | Farmer | 8(10.3) | 21(13.7) | 29(12.6) |
|  | Employee | 14(17.9) | 37(24.2) | 51(22.1) |
|  | Merchant | 10(12.8) | 16(10.5) | 26(11.3) |
| Family size | <4 | 37(47.4) | 61(39.9) | 98(42.4) |
|  | 4-6 | 32(41.0) | 70(45.8) | 102(44.2) |
|  | >6 | 9(11.5) | 22(14.4) | 31(13.4) |

Legend: \*others- single, divorced, widowed, Gov.-governmental, NGO-non governmental

### Medical illness and Obstetric complication factors

Among cases, 15(19.2%) of mothers had previous history of preterm birth, whereas eight (5.2%) of mothers in controls had previous history of preterm birth. About 24(30.8%) of cases and 15(9.8%) of controls had history of abortion during current pregnancy. Among cases, 11(14.1%) of mothers had not attended ANC, while only eight (5.2%) of mothers in controls group had no ANC attendance during current pregnancy. Among cases, about 13(16.7%) of mothers and 11(7.2%) of mothers in controls group had history of pregnancy induced hypertension. However, only 14(17.9%) of mothers among cases and 11(7.2%) of mothers in controls group had history of STI during current pregnancy. Among mothers participated in the study, about 168(72.7%) of mothers gave birth by spontaneously vaginal delivery, 38(16.5%) mothers gave birth by assisted instrumental delivery and 25(10.8%) of mothers gave birth by cesarean section delivery **(Table 2**)

**Table 2:** Medical illness and Obstetric Complication characteristics of mothers who gave birth in public hospitals of HGW zone, 2022 (n=231; case: 78 and control: 153)

| Medical and Obstetric complication factor |  | Case, n (%) | Control, n (%) | Total, n(%) |
| --- | --- | --- | --- | --- |
| Parity | ≤3 | 54(69.2) | 96(62.7) | 150(64.9) |
|  | 4-5 | 22(28.2) | 52(34.0) | 74(32.0) |
|  | >6 | 2(2.6) | 5(3.3) | 7(3.0) |
| Antenatal care | Yes | 67(85.9) | 145(94.8) | 212(91.8) |
|  | No | 11(14.1) | 8(5.2) | 19(8.2) |
|  | No visit | 11(14.1) | 8(5.2) | 19(8.2) |
| Number of ANC visit | Once | 10(12.8) | 8(5.2) | 18(7.8) |
|  | Twice | 28(35.9) | 32(20.9) | 60(26.0) |
|  | Three times | 20(25.6) | 54(35.3) | 74(32.0) |
|  | Four times | 9(11.5) | 51(33.3) | 60(26.0) |
| History of preterm birth | Yes | 16(20.5) | 12(7.8) | 28(12.1) |
|  | No | 62(79.5) | 141(92.2) | 203(87.9) |
| Family history of preterm birth | Yes | 12(15.4) | 23(15.0) | 35(15.2) |
|  | No | 66(84.6) | 130(85.0) | 196(84.8) |
| History abortion | Yes | 24(30.8) | 15(9.8) | 39(16.9) |
|  | No | 54(69.2) | 138(90.2) | 192(83.1) |
| Mode of delivery | Spontaneous | 61(78.2) | 107(69.9) | 168(72.7) |
|  | Instrumental | 11(14.1) | 27(17.6) | 38(16.5) |
|  | Cesarean section | 6(7.7) | 19(12.4) | 25(10.8) |
| Labor status | Spontaneously | 73(93.6) | 147(96.1) | 220(95.2) |
|  | Induced | 5(6.4) | 6(3.9) | 11(4.8) |
| History of Still birth | Yes | 5(6.4) | 15(9.8) | 20(8.7) |
|  | No | 73(93.6) | 138(90.2) | 211(91.3) |
| Antepartum hemorrhage | Yes | 10(12.8) | 14(9.2) | 24(10.4) |
|  | No | 68(87.2) | 139(90.8) | 207(89.6) |
| Pregnancy induced | Yes | 13(16.7) | 11(7.2) | 24(10.4) |
| Hypertension | No | 65(83.3) | 142(92.8) | 207(89.6) |
| Premature rupture of membrane | Yes | 12(15.4) | 6(3.9) | 18(7.8) |
|  | No | 66(84.6) | 147(96.1) | 213(92.2) |
| Birth interval (months) | <24 | 33(42.3) | 29(19.0) | 62(26.8) |
|  | >24 | 45 (57.7) | 124 (81.0) | 169 (73.2) |
| Sexually transmitted illness | Yes | 14(17.9) | 11(7.2) | 25(10.8) |
|  | No | 64(82.1) | 142(92.8) | 206(89.2) |
| History of malaria | Yes | 3 (3.8) | 2(1.3) | 5(2.2) |
|  | No | 75(96.2) | 151(98.7) | 226(97.8) |
| HIV test result | Positive | 1(1.3) | 2(1.3) | 3(1.3) |
|  | Negative | 77(98.7) | 151(98.7) | 228(98.7) |

Note: HIV: Human Immune Virus, ANC: Antenatal care

### Nutritional related characteristics

Regarding nutritional factors, among cases, 67(85.9%) of mothers had nutritional counseling whereas, 120(78.4%) of mothers among control group had no nutritional counseling during pregnancy. Among cases, about 19(24.4%) of mothers and 17(11.1%) of mothers among control group had history food taboo; whereas 26(33.3%) of cases and about 40(26.1%) of controls had MUAC <23cm during current pregnancy. Among cases, 21(26.9%) of mothers had anemia whereas 16(10.5%) of mothers in control group had anemia during their current pregnancy (**Table 3**).

**Table 3:** Nutritional related characteristics of mothers who gave birth in public hospitals of HGW zone, 2022 (n=231; case: 78 and control: 153)

| Nutritional Related Characteristics |  | Case, n (%) | Control, n(%) | Total, n(%) |
| --- | --- | --- | --- | --- |
| Nutritional counseling | Yes | 67(85.9) | 120(78.4) | 187(81.0) |
|  | No | 11(14.1) | 33 (21.6) | 44(19.0) |
| Meal frequency | $\leq 2$ | 12(15.4) | 21(13.7) | 33(14.3) |
|  | 3 | 37(47.4) | 72(47.1) | 109(47.2) |
| | $\geq 4$ | 29(37.2) | 60(39.2) | 89(38.5) |
| Additional meal | Yes | 20(25.6) | 21(13.7) | 41(17.7) |
|  | No | 58(74.4) | 132(86.3) | 190(82.3) |
| Food taboo | Yes | 24(30.8) | 19(12.4) | 43(18.6) |
|  | No | 54(69.2) | 134 (87.6) | 188(81.4) |
| Types of food taboo | None | 54(69.2) | 135(88.2) | 189(81.8) |
|  | Fruit and Vegetable | 0(0.0) | 3(16.7) | 3(7.1) |
|  | Egg and Meat | 17(70.8) | 11(61.1) | 28(66.7) |
|  | Milk and milk product | 7(29.2) | 4(22.2) | 11(26.2) |
| IFA supplement | Yes | 73(93.6) | 151(98.7) | 224(97.0) |
|  | No | 5(6.4) | 2(1.3) | 7(3.0) |
| Number IFA supplement | $\geq 90$tabs | 53(72.6) | 111(73.5) | 164(73.2) |
| | $< 90$tabs | 20(27.4) | 40(26.5) | 60(26.8) |
| Hgb level | $< 11$mg/dl | 21(26.9) | 16(10.5) | 37(16.0) |
| | $\geq 11$mg/dl | 57(73.1) | 137(89.5) | 194(84.0) |
| MUAC | $< 23$ | 26(33.3) | 40(26.1) | 66(28.6) |
| | $\geq 23$ | 52(66.7) | 113(73.9) | 165(71.4) |
| BMI | $< 18.5$ | 1(1.3) | 2(1.3) | 3(1.3) |
|  | 18.5-25 | 70(89.7) | 134(87.6) | 204(88.3) |
| | $> 25$ | 7(9.0) | 17(11.1) | 24(10.4) |

Note: IFA –Iron Sulphate and Folic Acid, Hgb-Hemoglobin, MUAC- Mid Upper Arm Circumference, BMI-Body Mass Index

### Lifestyle, Behavioral and Intimate Partner Violence Factors

Regarding behavior and lifestyle factor, about 47(60.3%) of cases and 57(37.3%) of controls had history of stressful life events during pregnancy. Among cases, about 14(17.9%) and 22(14.4%) of mothers in controls were alcohol users, whereas one (1.3%) in cases and two (1.3%) in controls had smoker husbands. However, only three (3.8%) of cases and one (0.7%) of mothers among controls had chewing khat during current pregnancy. On other hand, among cases 21(26.9%) and 19(12.4%) of control subjects had history of physical violence, whereas 21(26.9%) of cases and 26(17.0%) of controls had history of psychological violence during their current pregnancy (**Table 4**)

**Table 4:** Lifestyle, behavioral and intimate partner violence factors of mothers who gave birth in public hospitals of HGW zone, 2022 (n=231; case: 78 and control: 153).

| Lifestyle, behavioral and IPV related Characteristics |  | Case, n (%) | Control,n (%) | Total, n (%) |
| --- | --- | --- | --- | --- |
| Stressful life events | Yes | 47(60.3) | 57(37.3) | 104(45.0) |
|  | No | 31(39.7) | 96(62.7) | 127(55.0) |
| Reason for stressful life events | Hospitalization* | 29(61.7) | 29(51.8) | 58(56.3) |
|  | Family problem* | 18(38.3) | 27(48.2) | 45(43.7) |
| Drinking alcohol during pregnancy | Yes | 14(17.9) | 22(14.4) | 36(15.6) |
|  | No | 64(82.1) | 131(85.6) | 195(84.4) |
| Husband Smoking during pregnancy | Yes | 1(1.3) | 2(1.3) | 3(1.3) |
|  | No | 77(98.7) | 151(98.7) | 228(98.7) |
| Chewing Khat during pregnancy | Yes | 3(3.8) | 1(0.7) | 4(1.7) |
|  | No | 75(96.2) | 152(99.3) | 227(98.3) |
| Physical violence during pregnancy | Yes | 21(26.9) | 19(12.4) | 40(17.3) |
|  | No | 57(73.1) | 134(87.6) | 191(82.7) |
| Psychological violence during pregnancy | Yes | 21(26.9) | 26(17.0) | 47(20.3) |
|  | No | 57(73.1) | 127(83.0) | 184(79.7) |

Note:-\*Hospitalization: Illness, surgery, Hospitalization \* Family problem:-family conflict, family death

### Determinants of preterm birth

In the bivariable logistic regression analysis variables: residence, education level, Antenatal care attendance, history of Abortion, history of preterm birth, PROM, mode of delivery, anemia, Pregnancy induced hypertension, sexually transmitted illness, nutritional counseling, food taboo, stressful life events and physical violence were selected as candidate variable at p-value <0.25 for multivariable logistic regression. Multicollinearity was checked using variable inflation factor with result between 1.04 and 1.26. Hosmer-Lemeshow goodness fit test was checked model fitness with p-value indicated were 0.59. The result in multivariable logistic regression analysis showed that not attending antenatal care, history of abortion, premature rupture of the membrane, history of sexually transmitted illness and physical violence during pregnancy had statistical significant association with the preterm birth.

For instance, mothers who had antenatal care attendance during current pregnancy had 4.61 times higher odds of having preterm birth compared to mothers attended antenatal care (AOR=4.61, 95%CI; 1.54, 13.79). The odds of preterm birth among mothers experienced abortion prior to current pregnancy were four times higher compared to mothers with no history of abortion (AOR =3.88, 95% CI; 1.62, 9.30). Similarly, mothers who experienced premature rupture of membrane during current pregnancy had four times higher odds of preterm birth compared to mothers with no history of PROM (AOR=3.91, 95% CI; 1.15, 13.25).

Furthermore, the odds of preterm birth among mothers experienced sexually transmitted illness during current pregnancy were three folds higher compared to mothers without history of sexually transmitted illness (AOR=3.51, 95% CI; 1.26, 9.76). Moreover, the odds of preterm birth among mothers experienced physical violence during current pregnancy were three times higher compared to mothers with no history of physical violence (AOR=2.78, 95%CI; 1.19, 6.52) (**Table 5**)

**Table 5:** Determinants of preterm birth among mothers who gave birth in public hospitals of HGW zone, 2022 (n=231; case: 78 and control: 153).

| Variables |  | Case, n (%) | Control, n(%) | COR (95% CI) | AOR (95% CI) |
| --- | --- | --- | --- | --- | --- |
| Residence | Urban | 23 (29.5) | 69 (45.1) | 1.96 (1.10,3.51) | 0.73(0.35, 1.51) |
|  | Rural | 55(70.5) | 84(54.9) | 1 | 1 |
| Education level | No formal | 57(73.1) | 95(62.1) | 6.19(2.67,14.38) | 2.06(0.94, 4.55) |
|  | Formal | 21(26.7) | 58(37.9) | 1 | 1 |
| ANC visit | Yes | 67(85.9) | 145(94.8) | 1 | 1 |
|  | No | 11(14.1) | 8(5.2) | 2.98 (1.14 , 7.74) | **4.61 (1.54,13.79)\*\*\*** |
| History of Abortion | Yes | 24(30.8) | 15(9.8) | 4.09(1.99, 8.38) | **3.88(1.62,9.30)\*\*\*** |
|  | No | 54(69.2) | 138(90.2) | 1 | 1 |
| History of preterm | Yes | 16(20.5) | 12(7.8) | 2.84(1.26 ,6.43) | 2.49(0.93,6.67) |
|  | No | 62(79.5) | 141(92.2) | 1 | 1 |
| PROM | Yes | 12(15.4) | 6(3.9) | 4.46(1.60, 12.38) | **3.91(1.15,13.25)\*** |
|  | No | 66(84.6) | 147(96.1) | 1 | 1 |
| Mode of delivery | Spontaneous | 61(78.2) | 107(69.9) | 1 | 1 |
|  | Instrumental | 11(14.1) | 27(17.6) |  | 0.75(0.29,1.91) |
|  | C-section | 6(7.7) | 19(12.4) | 0.55(0.21,1.46) | 0.60(0.16,2.24) |
| STI | Yes | 14(17.9) | 11(7.2) | 2.82 (1.22, 6.56 ) | **3.51(1.26, 9.76)\*** |
|  | No | 64(82.1) | 142(92.8) | 1 | 1 |
| PIH | Yes | 13(16.7) | 11(7.2) | 4.455(1.60,12.38) | 2.26(0.83, 6.13) |
|  | No | 65(83.3) | 142(92.8) | 1 | 1 |
| hemoglobin level | <11mg/dl | 21(26.9) | 16(10.5) | 3.16 (1.54, 6.48) | 1.61(0.64, 4.09) |
|  | ≥11mg/dl | 57(73.1) | 137(89.5) | 1 | 1 |
| Stressful life events | Yes | 47(60.3) | 57(37.3) | 1.87 (1.07,3.24) | 1.33(0.68, 2.61) |
|  | No | 31(39.7) | 96(62.7) | 1 | 1 |
| Physical violence | Yes | 21(26.9) | 19(12.4) | 2.60 (1.30, 5.20) | **2.78(1.19, 6.52)\*** |
|  | No | 57(73.1) | 134(87.6) | 1 | 1 |
| Nutrition counseling | Yes | 67(85.9) | 120(78.4) | 1 | 1 |
|  | No | 11(14.1) | 33 (21.6) | 0.597(0.28,1.26) | 1.03(0.46, 2.51) |
| food taboo | Yes | 24(30.8) | 19(12.4) | 3.33 (1.68, 6.63) | 2.33 (0.94,4.55) |
|  | No | 54(69.2) | 134 (87.6) | 1 | 1 |

**Notes:** Reference = 1, \*P<0.05; \*\*P<0.01; \*\*\*P<0.001

## DISCUSSION

This study indicated that not attending ANC had higher risks of preterm birth. This result is consistent with study conducted in Pakistan(20), Ghana(21), Zambia(22), Amhara(23), Central Ethiopia(24) and Jimma(25). This could be because of mothers who had not attended ANC during pregnancy lacks awareness of obstetric complications and chance of identifying risk factors related to adverse birth outcome like preterm birth(26). This may decreases possible prevention and intervention mechanism, which in turn increases the risk of preterm birth. However, this study is not supported by the done in Silte, Ethiopia(27), which might be because of study period, study design, sample size and health care utilization in the area.

Similarly, in this study experiencing abortion on prior pregnancy had significant association with preterm birth. This finding is supported by the study conducted in Jimma (14), shire Tigray(12), Kenya(6) and Southern India(28). This might be because of the fact that surgical evacuation of uterus mechanically weakens the uterus and stretches the cervix, which predisposes mothers to give preterm birth in the consecutive pregnancies. Moreover, infection following the surgical procedure may extend to uterus resulting in intra-amniotic infection, which is idiopathic risk factor of preterm premature rupture of membrane, and result in preterm labor.

Likewise, this study signified that having history of premature rupture of membrane during pregnancy was found an independent predictor of preterm birth. This is agree with the study done in Ghana(21),Yemen(29),Amhara(30),Dilla(31) and Jimma(25). The possible justification might be activation of pro-inflammatory markers following premature rupture of membrane. The release of these cytokines stimulates the decidua layer of uterus and fetal membrane, which facilitate the release of some endogenous uterotonic hormones like prostaglandins and enzymes such as metalloproteinase; consequently, this may lead to an increased risk of enhanced uterine contraction, and finally, stimulates preterm labor(26, 32).

This study also indicated that there is significant association between history of sexually transmitted illness during pregnancy and preterm birth. This finding is consistent with study conducted in Sidama(33) and Pakistan(20). This might be because an ascending infection via vagina-cervix ultimately infects the amniotic sac and fluid, and this in turn ruptures the sac membrane and leaks amniotic fluid, which results in chorioamnionitis by the partial activation of systemic cytokine network and hematogenous dissemination, systemic infection and placental inflammatory response consecutively stimulate preterm labor(34). However, similar studies conducted in Somali, Ethiopia(13) revealed that mothers with the history of STI had no statistical association with Preterm birth. This might be due to difference in study design, study area, sample size and source population.

Furthermore, the result of this study revealed history of physical violence during pregnancy significantly associated with preterm birth. This finding is supported by the study conducted in Dilla(35) and Zimbabwe(36). The possible justification might be Violence could be committed against the victim physically, psychosocially, or sexually. Physical violence that involves the victim’s abdomen might cause placental injury and ruptures of the membranes. This initiates untimely uterine contraction resulting in preterm labor. On the otherhand, mothers who developed stress and depression following violence is found to have altered cell adhesion of molecules, pro-inflammatory cytokines, and raised C-reactive proteins. This could again lead to endothelial cell dysfunction and systemic inflammation to the placenta, which in turn triggers preterm labor resulting in preterm birth.

The limitation of the study includes, being institution–based study, institutional delivery is low. Thus, this study may lacks generalizability to entire population in the catchment areas. This study might also prone to recall bias, because of most question deals with the past information. However, to minimize potential recall bias, data for some important variables were crosschecked from medical records. This study might also be susceptible to social desirability bias due to face-to-face interview, which may underestimate reports from mothers. However, efforts have been made to minimize social desirability bias by choosing appropriate words in questioning and assuring confidentiality.

## Conclusion and Recommendation

The present study indicated that not attending antenatal care, history of abortion, premature rupture of membranes, sexually transmitted illness and physical violence during current pregnancy had independent statistical association with preterm births. Furthermore, focused antenatal care service and proper risk managements may minimize the identified factors.

Therefore, Federal ministry of health and non-governmental organization should create public awareness through public health communication program regarding to determinants of preterm birth, particularly, antenatal care service, sexually transmitted illness, abortion and physical violence. Moreover, essential maternal and newborn intervention program, particularly, Neonatal Intensive Care Unit service should be implemented in primary health facilities. Similarly, healthcare provider and public hospitals should conduct screening of pregnant mothers, identify obstetric complications; encourage pregnant mothers to attend packages of focused ANC service, prevent abortion and conduct safe abortion care service.

In addition, community based health care providers and health extension workers should counsel pregnant mother focusing harmful consequences of identified determinants of pregnancy like abortion, sexually transmitted illness and physical violence. Lastly, future researcher should conduct stronger longitudinal and community-based studies to create causal relationship.

## Data Availability

check the manuscript accordingly

## Acknowledgements

First and for most, I would like to thank Wallaga University, Institute of health sciences, department of public health for its academic instructions and facilitate to conduct this research work. Next, I would like to express my deepest gratitude and appreciation to my advisors Mr. Emiru Merdassa and Mr. Bizuneh Wakuma from Institute of health Sciences, Wallaga University for providing me insights, comments, valuable assistance and unreserved guidance throughout the completion of this thesis work. I would like to extend my heartfelt thanks to all my colleagues and staff in public hospitals of Horro Guduru Wallaga zone, data collectors, supervisors and study participants for their active participation in providing necessary information during data collection.

## Author Contributions

All authors were actively involved in the planning and design of the study Warkisa Bayisa Duresa was principal writer of the paper, and collected and analyzed the data. Emiru Merdassa Atomsa and Bizuneh Wakuma helped in interpretation of the data and writing of the paper. Worku Etafa took part in the study design and developments of this document. All authors undertook all statistical analyses and wrote the original draft of the paper. Warkisa Bayisa Duresa, Emiru Merdassa Atomsa, Bizuneh Wakuma and Worku Etafa were contributed to the final version of the paper and act as guarantors.

## REFERENCES

1. Quinn J-A, Munoz FM, Gonik B, Frau L, Cutland C, Mallett-Moore T, et al. Preterm birth: Case definition & guidelines for data collection, analysis, and presentation of immunisation safety data. Vaccine. 2016;34(49):6047–56.

2. Walani SR. Global burden of preterm birth. International Journal of Gynecology & Obstetrics. 2020;150(1):31–3.

3. Bick D. Born too soon: the global issue of preterm birth. Midwifery. 2012;28(4):341–2.

4. Chawanpaiboon S, Vogel JP, Moller A-B, Lumbiganon P, Petzold M, Hogan D, et al. Global, regional, and national estimates of levels of preterm birth in 2014: a systematic review and modelling analysis. The Lancet Global Health. 2019;7(1):e37–e46.

5. van den Broek NR, Jean-Baptiste R, Neilson JP. Factors associated with preterm, early preterm and late preterm birth in Malawi. PloS one. 2014;9(3):e90128.

6. Okube OT, Sambu LM. Determinants of preterm birth at the postnatal ward of Kenyatta National Hospital, Nairobi, Kenya. Open Journal of Obstetrics and Gynecology. 2017;7(09):973.

7. Preemie–SCALE E. Ethiopia: profile of preterm and low birth weight prevention and care. 2019.

8. Gebreslasie K. Preterm birth and associated factors among mothers who gave birth in Gondar town health institutions. Advances in Nursing. 2016;2016.

9. Cherie N, Mebratu A. Adverse Birth Out Comes and Associated Factors among Delivered Mothers in Dessie Referral Hospital. North East Ethiopia. 2018:1–6.

10. Abdo RA, Halil HM, Muhammed MA, Karebo MS. Magnitude of Preterm Birth and Its Associated Factors: A Cross-Sectional Study at Butajira Hospital, Southern Nations, Nationalities, and People’s Region, Ethiopia. International Journal of Pediatrics. 2020;2020.

11. Berhe T, Gebreyesus H, Desta H. Determinants of preterm birth among mothers delivered in Central Zone Hospitals, Tigray, Northern Ethiopia. BMC Res Notes. 2019;12(1):266.

12. Kelkay B, Omer A, Teferi Y, Moges Y. Factors associated with singleton preterm birth in Shire Suhul general hospital, northern Ethiopia, 2018. Journal of pregnancy. 2019;2019.

13. Muhumed II, Kebira JY, Mabalhin MO. Preterm Birth and Associated Factors Among Mothers Who Gave Birth in Fafen Zone Public Hospitals, Somali Regional State, Eastern Ethiopia. Research and Reports in Neonatology. 2021;11:23–33.

14. Bekele I, Demeke T, Dugna K. Prevalence of preterm birth and its associated factors among mothers delivered in Jimma university specialized teaching and referral hospital, Jimma Zone, Oromia Regional State, South West Ethiopia. J Women’s Health Care. 2017;6(1):1–10.

15. Blencowe H, Cousens S, Chou D, Oestergaard M, Say L, Moller AB, et al. Born too soon: the global epidemiology of 15 million preterm births. Reprod Health. 2013;10 Suppl 1(Suppl 1):S2.

16. Barfield WD. Public Health Implications of Very Preterm Birth. Clin Perinatol. 2018;45(3):565–77.

17. Goldenberg RL, Rouse DJ. Prevention of premature birth. New England Journal of Medicine. 1998;339(5):313–20.

18. Hoeltl A, Brandtweiner R, Bates R, Berger T. The interactions of sustainable development goals: The case of urban informal settlements in Ethiopia. International Journal of Sustainable Development and Planning. 2020;15(3):287–94.

19. Bekele A, Mussema Y, Tadesse Y, Taylor ME. Reaching every newborn: delivering an integrated maternal and newborn health care package. Ethiopian Medical Journal. 2019(3).

20. Hanif A, Ashraf T, Pervaiz MK, Guler N. Prevalence and risk factors of preterm birth in Pakistan. JPMA The Journal of the Pakistan Medical Association. 2020;70(4):577–82.

21. Aseidu EK, Bandoh DA, Ameme DK, Nortey P, Akweongo P, Sackey SO, et al. Obstetric determinants of preterm delivery in a regional hospital, Accra, Ghana 2016. BMC Pregnancy Childbirth. 2019;19(1):248.

22. Mukosha M, Jacobs C, Musonda P, Zulu JM, Masaku S, Nkwemu C, et al. Determinants of Preterm Births at a National Hospital in Zambia: Application of Partial Proportional Odds Model. Obstetrics and Gynecology Research. 2021;4:117–30.

23. Woday A, Muluneh MD, Sherif S. Determinants of preterm birth among mothers who gave birth at public hospitals in the Amhara region, Ethiopia: A casecontrol study. PloS one. 2019;14(11):e0225060.

24. Deriba BS, Ayalew AF, Gebru AA. Determinants of preterm birth in public hospitals in central Ethiopia: an unmatched case-control study. F1000Research. 2021;10(773):773.

25. Abaraya M, Seid SS, Ibro SA. Determinants of preterm birth at Jimma university medical center, Southwest Ethiopia. Pediatric health, medicine and therapeutics. 2018;9:101.

26. Abadiga M, Wakuma B, Oluma A, Fekadu G, Hiko N, Mosisa G. Determinants of preterm birth among women delivered in public hospitals of Western Ethiopia, 2020: Unmatched case-control study. Plos one. 2021;16(1):e0245825.

27. Hassen JA, Handiso MN, Admassu BW. Predictors of Preterm Birth among Mothers Who Gave Birth in Silte Zone Public Hospitals, Southern Ethiopia. J Pregnancy. 2021;2021:1706713.

28. Rao D, Kumar S, Mohanraj R, Frey S, Manhart LE, Kaysen DL. The impact of domestic violence and depressive symptoms on preterm birth in South India. Social psychiatry and psychiatric epidemiology. 2016;51(2):225–32.

29. Dahman HAB. Risk factors associated with preterm birth: a retrospective study in Mukalla Maternity and Childhood Hospital, Hadhramout Coast/Yemen. Sudanese Journal of Paediatrics. 2020;20(2):99.

30. Adugna DG, Oumar M, Adugna A. Premature Babies and Associated Factors Among Births in Referral Hospitals of Amhara Region, Ethiopia: a Cross-sectional Study. 2021.

31. Zewde A, Gebremichael B, Tesfaye T, Sibhat MM, Bimer K. Risk factors of preterm birth among newborns delivered in public hospitals of Southern Ethiopia: A case-control study. 2022.

32. Sari IM, Adisasmita AC, Prasetyo S, Amelia D, Purnamasari R. Effect of premature rupture of membranes on preterm labor: a case-control study in Cilegon, Indonesia. Epidemiology and health. 2020;42.

33. Sifer S, Kedir B, Demisse G, Sisay Y. Determinants of preterm birth in neonatal intensive care units at public hospitals in Sidama zone, South East Ethiopia; case control study. J Pediatr Neonatal Care. 2019;9(6):180–6.

34. Gao R, Liu B, Yang W, Wu Y, Wang B, Santillan MK, et al. Association of maternal sexually transmitted infections with risk of preterm birth in the United States. JAMA network open. 2021;4(11):e2133413–e.

35. Zewde GT. Preterm Birth and Associated Factors Among Mother Who Gave Birth in Public Health Hospitals in Harar Town Eastern Ethiopia 2019. OSP Journal of Health Care and Medicine. 2020;1(1):1–3.

36. Yaya S, Odusina EK, Adjei NK, Uthman OA. Association between intimate partner violence during pregnancy and risk of preterm birth. BMC public health. 2021;21(1):1–9.

